# Two New Models for Epidemics with Application to the COVID-19 Pandemic in the United States, Italy, and the United Kingdom

**DOI:** 10.1101/2020.07.13.20152686

**Authors:** David H. Roberts

## Abstract

The Distributed Logistic Model and the Adaptive Logistic Model of epidemics are formulated and used to study the course of cases and deaths during the COVID-19 pandemic. The distributed model is designed to account for a spread of initiation times of hot spots across a country; it does especially well at capturing the initial and linear phases of epidemics. The adaptive model accounts for the development of social mitigation factors, and does especially well at capturing the declining phases of epidemics. The historical data for the U.S., Italy, and the U.K. are analyzed in detail. The parameters of the fits to the two models provide complementary information about the pandemic. The initial infection rate constant was *r*_0_ ≃ 0.29 day^−1^ for each country, and the effective infection rate constants evolved with time in essentially the same way for each. This suggests that mitigation effects were equally effective in all three countries. Analysis with the distributed model suggests that it took somewhat different times *T* for the epidemic to spread across each country, with *T* (US)≃ 50 days significantly greater than the *T* ‘s of Italy or the U.K. The mortality ratio in the U.S. was about 0.061 while in Italy and U.K. it was much larger at about 0.15.

## 1. Introduction

Currently a pandemic of the novel coronavirus (SARS-CoV-2) is overspreading the world. The United States has been particularly impacted by the disease it causes, COVID-19. Epidemiologists have worked to describe adequately this pandemic and to predict its future course. This is a complex undertaking, involving the mathematics of epidemiology and a vast amount of uncertain data that serve as input to the models (see, for example, CCD 2020; Fivethirtyeight 2020).

This paper derives two new epidemiological models, the *Distributed Logistic Model* and the *Adaptive Logistic Model*, and uses them to study the course of the COVID-19 pandemic in the U.S., Italy, and the U.K. The goal of each model is to extract different information about the progress of each epidemic; they are not merely *ad hoc* parameterizations designed to fit the data.

The plan of this paper is as follows: Section 2 reviews standard epidemiological approaches to the study of the COVID-19 pandemic, section 3 introduces the basic logistic model, section 4 derives the Distributed Logistic Model, section 5 derives the Adaptive Logistic Model, section 6 uses these two models to extract new information about each epidemic, and section 7 discusses the results.

## 2. Approaches to the Study of Epidemics

### 2.1. The SIR and Other Models for Epidemics

The study of the spread of disease through a population is a highly mathematical enterprise involving the solution of multiple coupled ordinary differential equations (see, for example, Hethcote 2000). The fundamental set of equations are the Susceptible-Infected-Recovered/Removed (“SIR”) equations (Kermack & McKendrick 1927) and their extensions and generalizations (Hethcote 2000 and references therein). These equations describe the flow of individuals among three or more classes using a series of coupled rate equations. In some cases (Lavrova *et al*. 2017) they can be reduced to the single logistic differential equation (Verhulst (1838), Equation 1 below).

### 2.2. Applications of the SIR Equations to the 2020 COVID-19 Pandemic

Numerous authors have applied the SIR equations or their generalizations to the study of the 2020 COVID-19 pandemic. Huang *et al*. (2020) used the standard SIR model to examine the COVID-19 pandemic in China, Bulgaria, Costa Rica, Faroe Islands, French Guiana, Maldives, Malta, Martinique, and Republic of Moldova. Chen *et al*. (2020) used a time-dependent SIR model to study the outbreaks in China, South Korea, Italy, and Iran. They concluded that various approaches to social distancing could lead to a reduction in the basic reproductive number *R*_0_. Zhao and Chen (2020) applied a Susceptible, Un-quarantined infected, Quarantined infected, Confirmed infected (SUQC) model to study several cities and regions of China. They extracted the reproductive numbers *R* and other parameters, and predicted the course of the epidemics. Wangping *et al*. (2020) used an extended susceptible-infected-removed (eSIR) model to study the epidemic in Italy and compare it to that in China.

The American Hospital Association keeps an updated compendium of the models being used to track the COVID-19 pandemic (AHA 2020), as does the Centers for Disease Control and Prevention (CDC 2020). Among the goals of these studies has been to predict the future course of the pandemic. The results have varied widely, and have not engendered a great deal of confidence in this enterprise (e.g., Best & Boice (2020), CDC (2020)).

### 2.3. Applications of the Logistic Equation and Its Generalizations

Other authors have used the logistic equation (Equation 1 below) or its generalizations to analyze the epidemics in various countries. Shen (2020) used the logistic equation to study the outbreaks in ten Chinese municipalities and nine western countries. They concluded that there is potential for this model to contribute to better public health policy in combatting COVID-19. This study also outlined certain challenges in modeling and their implications for the results. Chen, Chen, & Chen (2020) studied the epidemic in the United States using a five-parameter logistic model,

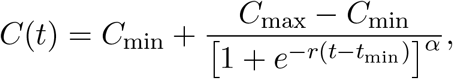

where *C*(*t*) is the population of infected individuals. This is a generalization of the solution to the logistic equation given in Equation 2 below. Here *t*_min_ and *α* are *ad hoc* parameters. These authors demonstrated the ability of this model to fit the U.S data. Wu *et al*. (2020) studied China and eight European countries using either a generalized logistic model

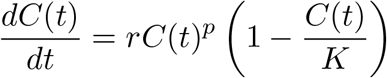

or a generalized Richards model

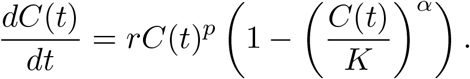

Here *p* and *α* are again *ad hoc* parameters; while they can be optimized to produce excellent fits to the data, and thus to forecast the future, they cannot be interpreted physically. Batista (2020) used the logistic growth model for estimation of the final sizes and peak times of the coronavirus epidemic in China, South Korea, and the rest of the world.

Except for Wu *et al*. (2020), these models provided only modestly good fits to the data. In no case could the added parameters be interpreted physically. The sections below show how this situation can be improved by two well motivated modifications of the logistic equation.

### 2.4. Approach in This Paper

The current paper takes a different tack toward providing a mathematical description of epidemics. The procedure will be to make two separate modifications to the logistic equation, changes that represent two effects that are neglected in derivation of the basic logistic equation. The goal is to use the data to determine the values of the new parameters that are introduced, and then to interpret them in terms of the course of the spread of the disease and of the response of humanity to the pandemic.

The history of the epidemic in each country is encapsulated in a time series of cases of infections and a time series of deaths. The sections below analyze these data using the basic logistic equation and the two modifications. The reader should note that the author is a physicist not an epidemiologist, and that his approach will be intrinsically that of his discipline. However, the results of the analyses will be connected to the real world environment of the pandemic.

## 3. Basic Logistic Model

### 3.1. Derivation

The simplest model for the evolution of an epidemic is based on the logistic differential equation. This describes an epidemic that begins with a small number *f*_0_ of infected individuals at a single time and place, and subsequently spreads through a population. The motivation for this model is as follows. If the population of infected individuals (i.e., the total number of cases of disease) as a function of time is *f* (*t*), simple exponential growth with growth rate constant *r* is determined by the differential equation

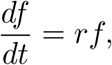

with the growing exponential solution

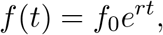

where *f*_0_ = *f* (0). That is what happens with an unlimited pool of subjects. However, for a finite number of subjects *K*, as the population of infected individuals grows the number of subjects available to be infected is reduced by the factor (1 − *f/K*); taking this into account leads to the *logistic differential equation*,^2^

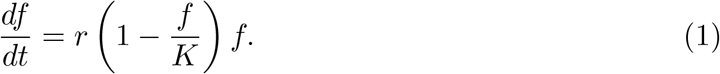

The solution of this equation is well known to be

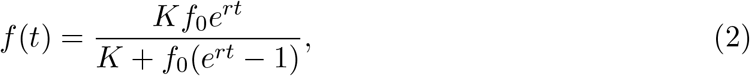

which satisfies the conditions *f* (0) = *f*_0_ and *f* (∞) = *K*. The time course described by Eqn. 2 is the familiar S curve used to describe bacterial growth and other rate limited phenomena (see Fig. 1 for examples). The history of the total number of cases of disease can be fitted by this equation to determine the parameters *r* and *K*, each of which has a well-defined meaning. In particular, *K* represents a prediction of the endpoint of the epidemic.

**Fig. 1.**
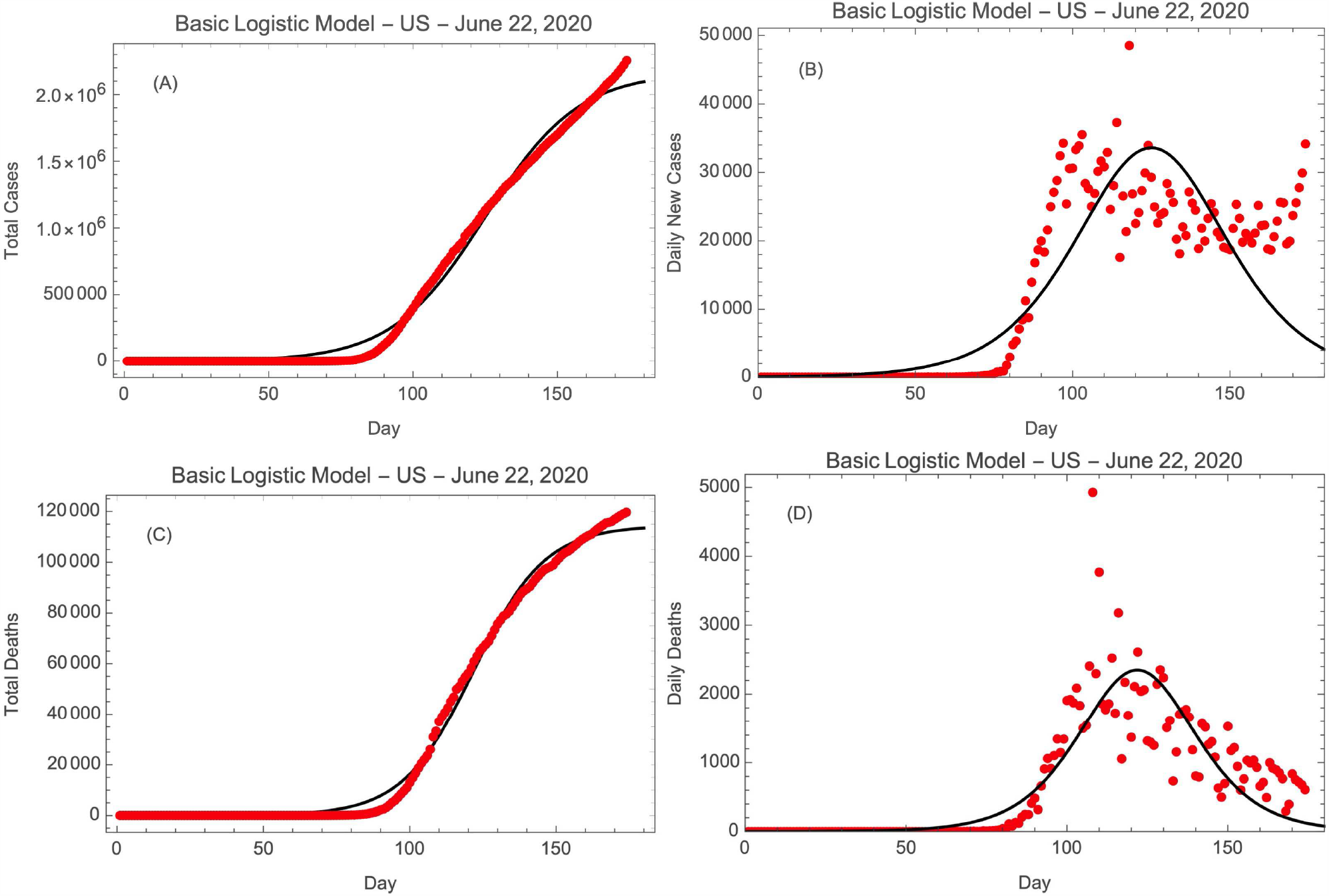
BLM fits to (A) total cases, (B) daily cases, (C) total deaths, and (D) daily deaths in the U.S. The points are the data and the curves are the fits to Eqns. 2 & 3

**Fig. 2.**
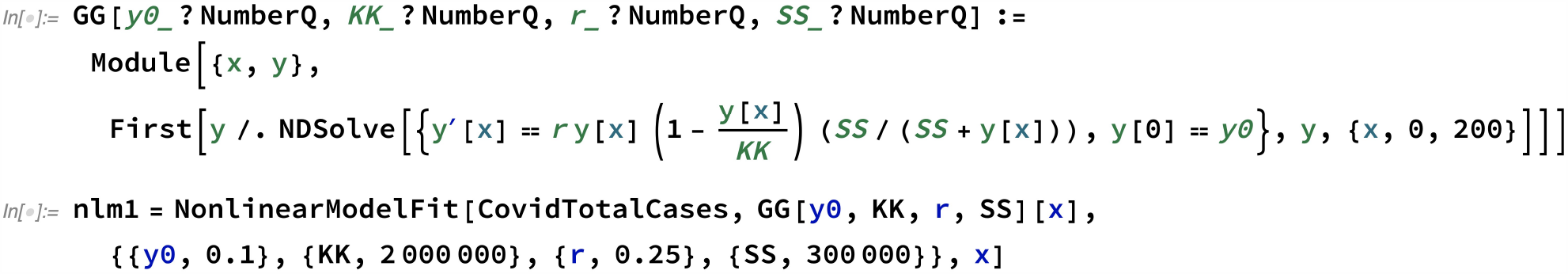
Mathematica™ code for optimization of a theoretical expression that can only be determined by numerical solution of a differential equation.

An analysis of the total number of cases as a function of time with *f* (*t*) is not the only way to compare model and data. Instead one can examine the number of new cases per day as a function of time;^3^ in the model this is the time derivative of *f* (*t*). This is found as a function of time by substituting Eqn. 2 into Eqn. 1, giving

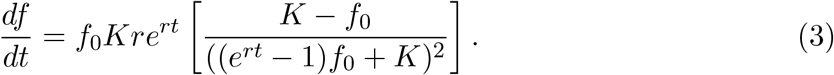

In either case, for the logistic model the three parameters to be adjusted are *f*_0_, *K*, and *r*. For this basic logistic model the total counts and the daily counts are fit independently. The fits to the U.S. data on number of cases and number of deaths^4^ are shown in Fig. 1. The basic logistic model will be referred to as the BLM.

## 4. The Distributed Logistic Model

### 4.1. Derivation

In reality the COVID-19 epidemic in the US is a series of epidemics centered at various places distributed across the country and beginning over a range of times. This can be accounted for this by averaging a series of logistic distributions over a range of starting times *t*_i_ spanning a total time *T* ; the appropriate function to be averaged is

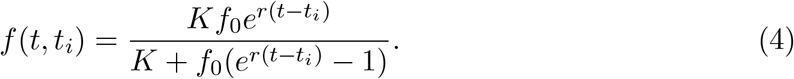

To account for a total range of time *T* over which the various hot spots of the epidemic are distributed, one integrates Eqn. 4 over 0 ≤ *t*_i_ ≤ *T* and divides by *T*. The result is the *distributed logistic function*

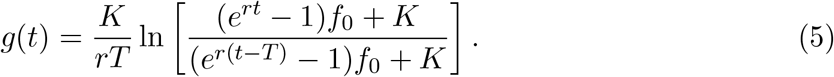

Note that

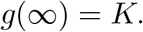

The differential distribution is derived from Eqn. 5,

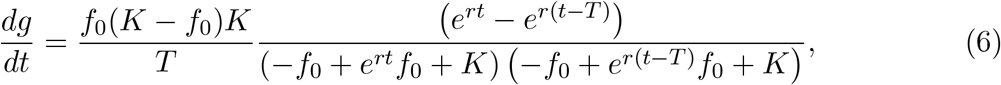

and is used to model the daily data. For the Distributed Logistic Model the four parameters to be adjusted are *f*_0_, *K, r*, and *T* ; the total number and the daily numbers are fit independently. The fits to the U.S. data on cases of disease and on deaths are shown in Fig. 3 below. The Distributed Logistic Model will be referred to as the DLM.^5^

**Fig. 3.**
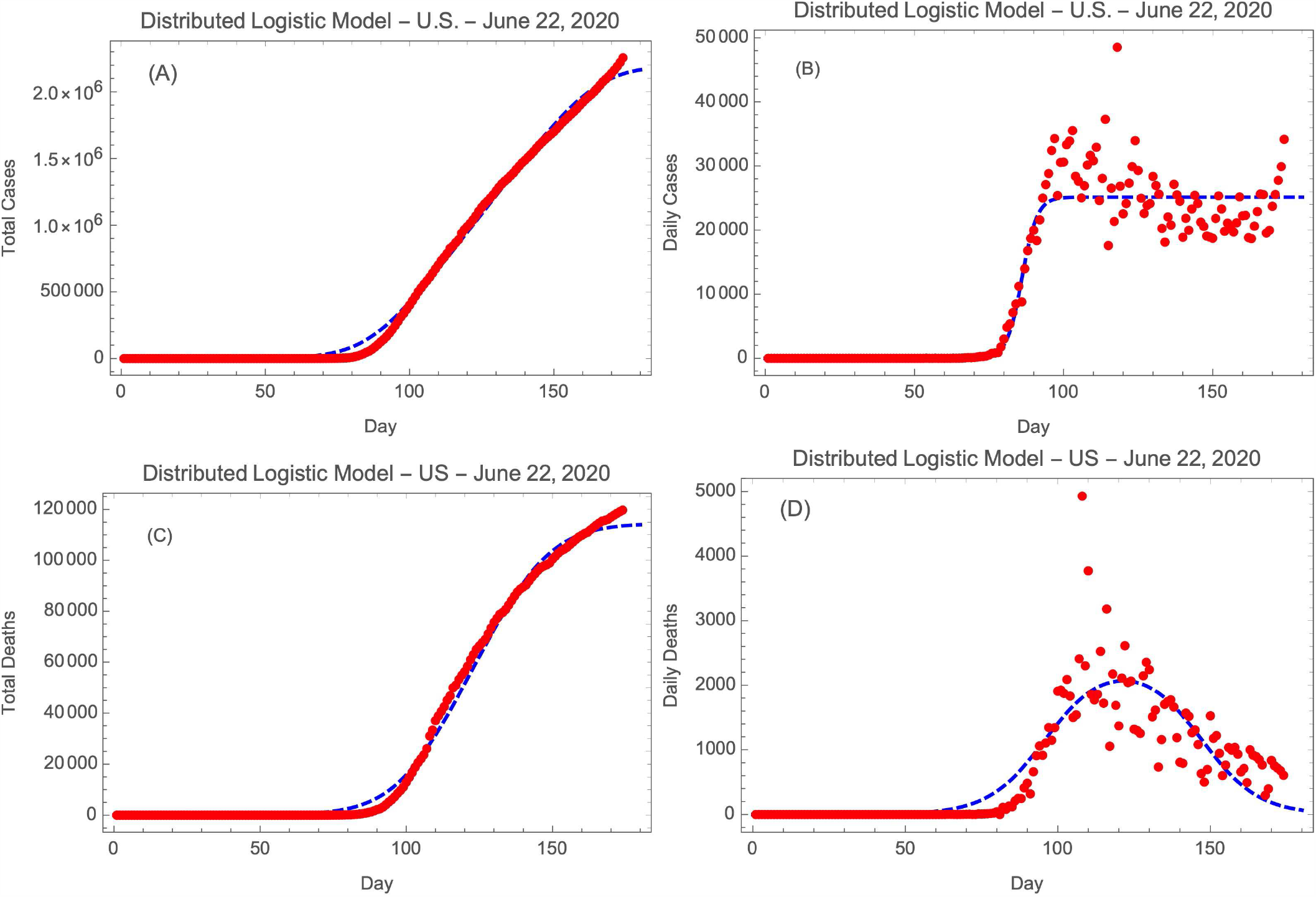
DLM fits to (A) total cases, (B) daily cases, (C) total deaths, and (D) daily deaths in U.S. The points are the data and the curves are the fits to Eqns. 5 & 6.

## 5. The Adaptive Logistic Model

### 5.1. Derivation

During the course of the COVID-19 pandemic, countries across the world introduced a number of measures to reduce the spread of the disease, including quarantining, masking, social distancing, and contact tracing and isolating. As the pandemic progressed these measures tended to become more effective. Their effect was to make the average infected individual progressively less likely to infect another person. A factor that reduces the probability of transmission as the pandemic progresses can be introduced into Eqn. 1 to account for this effect. A simple way to do this is to modify the infection rate constant *r* by a function that decreases as the infected population increases. After some experimentation,^6^ the function *S/*(*S*+*g*) = 1*/*(1+*g/S*), where *S* is a dimensionless parameter to be determined from the data, was chosen, leading to the *adaptive logistic equation*^7^

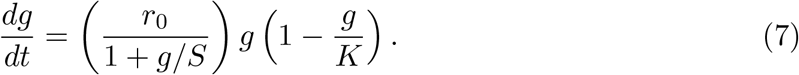

Here *r*_0_ is the infection rate constant at the beginning of the epidemic (*g* ≪ *S*). When *g* reaches *S* the initial infection rate constant is halved, so in that sense *S* is its half-life. The function

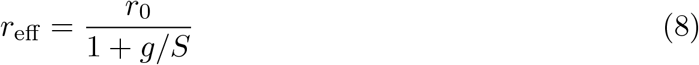

will be referred to as the *effective infection rate constant*; as the population *g* increases *r*_eff_ decreases, so r_eff_ is a function of time. The Adaptive Logistic Model fits to the U.S. data are shown in Fig. 4. The Adaptive Logistic Model will be referred to as the ALM.

**Fig. 4.**
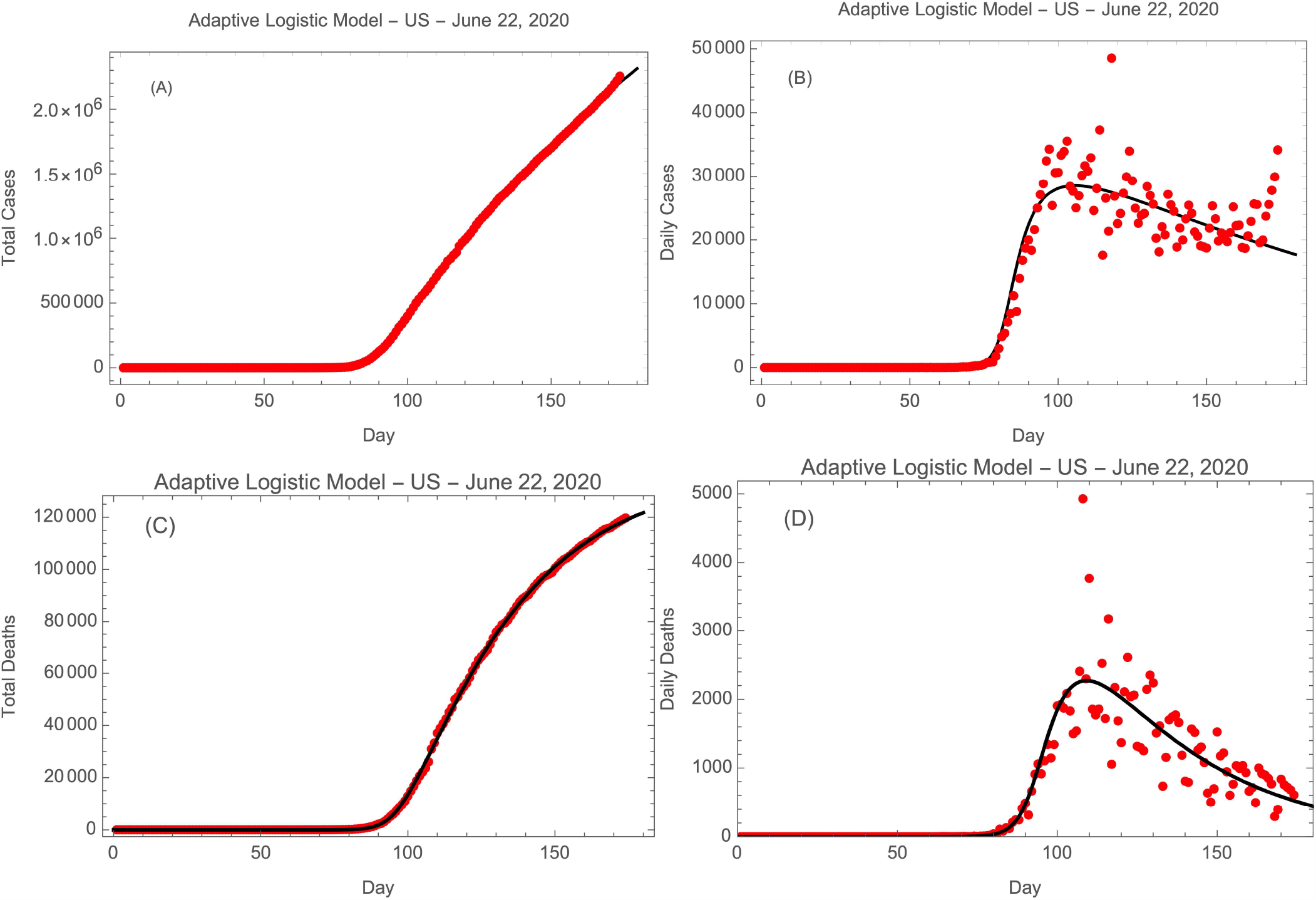
ALM fits to (A) total cases, (B) daily cases, (C) total deaths, and (D) daily deaths in the U.S. The points are the data, the total numbers curves are the fits to Eqn. 7, whereas the daily numbers curves are their derivatives.

### 5.2. Method of Solution

For the BLM and the DLM the parameters were estimated from the data by use of non-linear least squares minimization of the differences between the data and the prediction of Eq. 1, 3, 5, or 6 using the NMinimize function in Mathematica™. A closed form solution to Eqn. 7 could not be found, so it was solved and fit to the data numerically. This was done with Mathematica™ code whose key parts are shown in Fig. 2. In this example, the matrix CovidTotalCases contains the data.^8^ To determine the predicted distributions of the number of daily cases or deaths one takes the numerical derivatives of the fits to *g*(*t*). It must be emphasized that this does not fit the daily numbers, it merely compares them with the expectations derived from the fits to the total numbers.^9^ The Adaptive Logistic Model fits to the U.S. data are shown in Figure 4. The Adaptive Logistic Model will be referred to as the ALM.

## 6. The Analyses of the Pandemic

The logistic equations are intended to describe the time behavior of the number of cases of disease. However, if the mortality ratio is roughly time independent for a given country, the same functions will describe the history of deaths;^10^ thus both cases and deaths are analyzed using Equations 2, 3, 5, 6, & 7. The BLM was applied only to the U.S. data.

The data were taken from ECDPC (2020); they span January 31, 2019 through June 22, 2020 (174 days).

### 6.1. The Epidemic in the United States

Figures 1, 3, & 4 shows the fits of the BLM, the DLM, and the ALM to the histories of cases and deaths in the U.S. The parameters of the fits to the DLM and the ALM are displayed in Tables 1 & 2.

**Table 1.** Parameters of the Distributed Logistic Model Fits.

| Parameter | U.S. Cases | U.S. Deaths | Italy Cases | Italy Deaths | U.K. Cases | U.K. Deaths |
| --- | --- | --- | --- | --- | --- | --- |
| $N_{\text{data points}}$ | 174 | 174 | 174 | 174 | 174 | 174 |
| $N_{\text{tot}}$ | 2,255,119 | 119,719 | 238,275 | 34,610 | 303,110 | 42,589 |
| $K^a$ | $2.6 \times 10^6$ | $1.2 \times 10^5$ | $2.3 \times 10^5$ | $3.3 \times 10^4$ | $3.0 \times 10^5$ | $4.0 \times 10^4$ |
| $r \text{ (day}^{-1}\text{)}^a$ | 0.278 | 0.106 | 0.101 | 0.116 | 0.113 | 0.100 |
| $T \text{ (days)}^a$ | 103. | 51. | 35. | 37. | 53. | 29. |
| $M \text{ (Mortality Ratio)}^b$ | ... | 0.0531 | ... | 0.145 | ... | 0.141 |
| $RMS_{\text{daily}}$ | 4600. | 430. | 590. | 85. | 680. | 150. |
| $RMS_{\text{daily}} \times M^c$ | 240. | ... | 41. | ... | 96. | ... |
| $\chi^2_{\text{daily}}/\text{d.o.f.}$ | 1300. | 230. | 4450. | 170. | 370. | 130. |
| $\chi^2_{\text{daily}}/\text{d.o.f.} \times M^c$ | 73. | ... | 65. | ... | 14. | ... |

<sup>a</sup>These entries were determined from the daily distributions.
<sup>b</sup>Total deaths divided by total cases.
<sup>c</sup>Scaled by mean mortality ratio to compare cases data with deaths data.

**Table 2.**
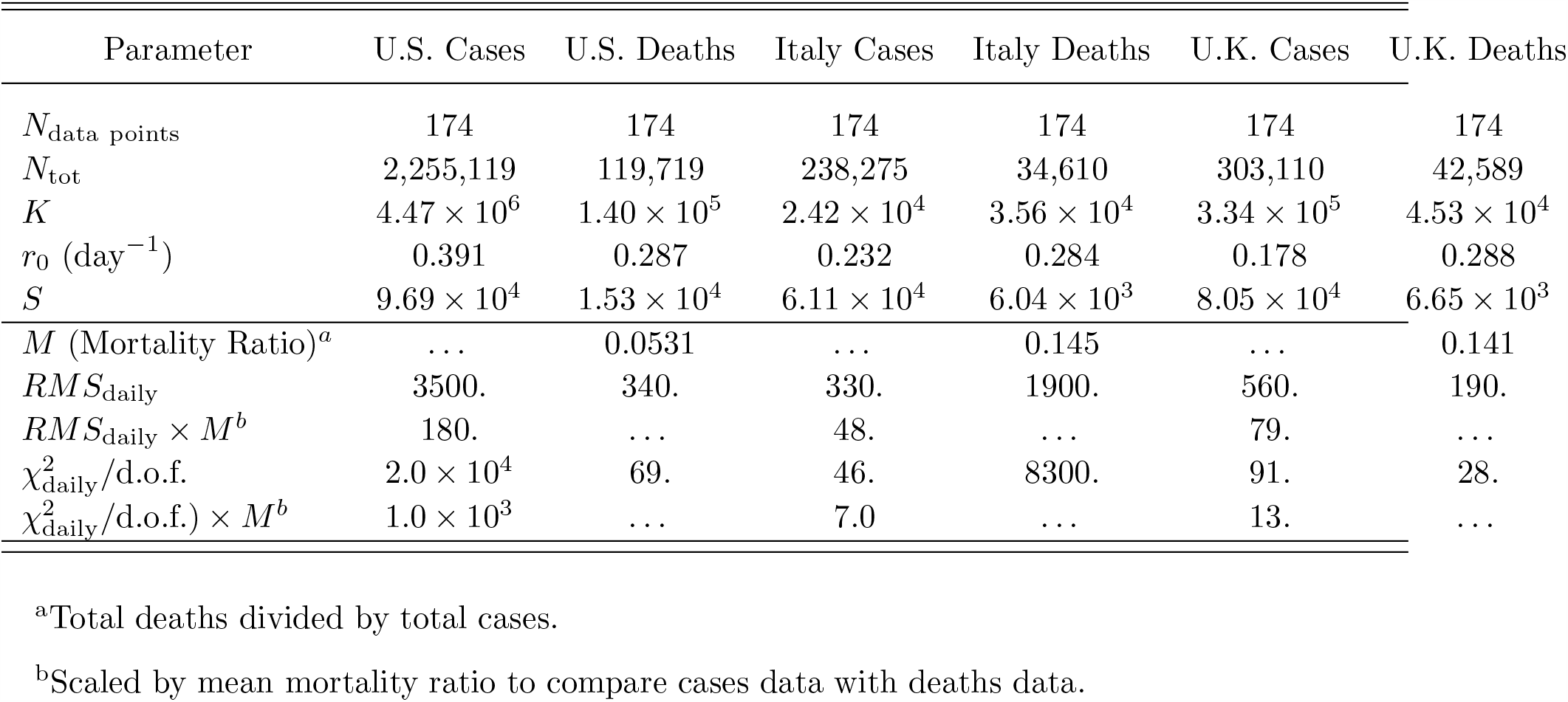
Parameters of the Adaptive Logistic Model Fits.

| Parameter | U.S. Cases | U.S. Deaths | Italy Cases | Italy Deaths | U.K. Cases | U.K. Deaths |
| --- | --- | --- | --- | --- | --- | --- |
| $N_{\text{data points}}$ | 174 | 174 | 174 | 174 | 174 | 174 |
| $N_{\text{tot}}$ | 2,255,119 | 119,719 | 238,275 | 34,610 | 303,110 | 42,589 |
| $K$ | $4.47 \times 10^6$ | $1.40 \times 10^5$ | $2.42 \times 10^4$ | $3.56 \times 10^4$ | $3.34 \times 10^5$ | $4.53 \times 10^4$ |
| $r_0 \text{ (day}^{-1}\text{)}$ | 0.391 | 0.287 | 0.232 | 0.284 | 0.178 | 0.288 |
| $S$ | $9.69 \times 10^4$ | $1.53 \times 10^4$ | $6.11 \times 10^4$ | $6.04 \times 10^3$ | $8.05 \times 10^4$ | $6.65 \times 10^3$ |
| $M \text{ (Mortality Ratio)}^a$ | ... | 0.0531 | ... | 0.145 | ... | 0.141 |
| $RMS_{\text{daily}}$ | 3500. | 340. | 330. | 1900. | 560. | 190. |
| $RMS_{\text{daily}} \times M^b$ | 180. | ... | 48. | ... | 79. | ... |
| $\chi^2_{\text{daily}}/\text{d.o.f.}$ | $2.0 \times 10^4$ | 69. | 46. | 8300. | 91. | 28. |
| $\chi^2_{\text{daily}}/\text{d.o.f.} \times M^b$ | $1.0 \times 10^3$ | ... | 7.0 | ... | 13. | ... |

<sup>a</sup>Total deaths divided by total cases.
<sup>b</sup>Scaled by mean mortality ratio to compare cases data with deaths data.

### 6.2. Comparison of the Models

Figures 1, 3, & 4 show the fits of the three logistic models to the same four U.S. data sets. It is apparent from these figures that the BLM does poorly at fitting any of the data sets, and it will not be considered further.^11^ It is for this reason that the other two models were derived; the motivations for their specific forms are given with their derivations. Now the question is, which of these two new models provides a superior representative of the data? Figure 5 is a comparison of the DLM and ALM fits to the daily U.S. cases data. Both models are successful at fitting the exponential rise and subsequent linear phase, while the ADM is more successful at fitting the declining phase of the epidemic. The parameter *T* in the DLM tells us the length of time it has taken the epidemic to spread across each country, while the parameter *S* in the ALM contains information on the time history of the effective infection rate constant.

**Fig. 5.**
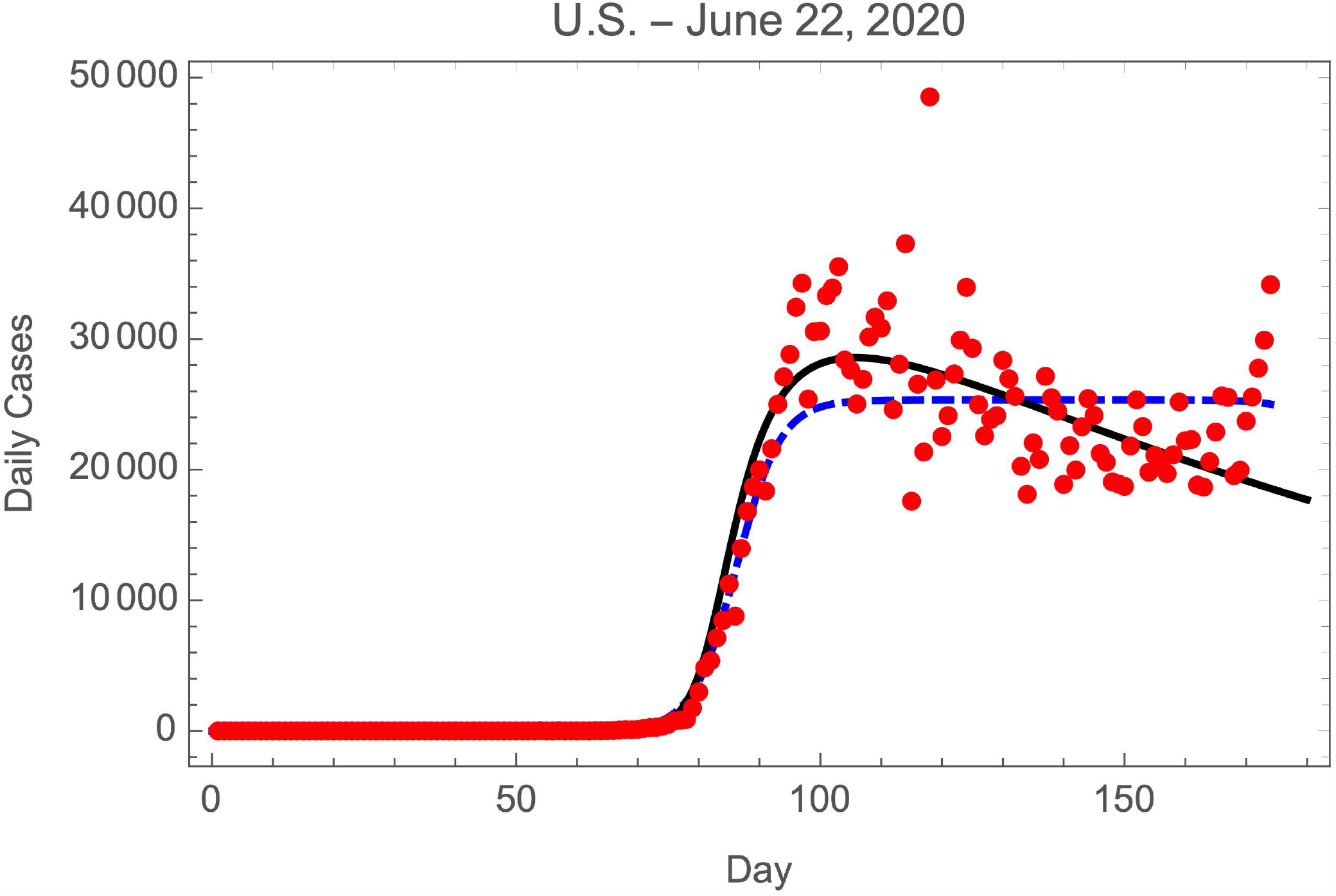
Comparison of the DLM & ALM fits to daily cases data in the U.S. The points are the data, the fit to the DLM is the dashed blue curve, and the fit to the ALM is the black curve.

Two measures of the qualities of the fits are shown in Tables 1 & 2, the scaled^12^ root-mean-squared deviation for the daily data (RMS_daily_) and the scaled chi-squared per degree of freedom^13^ for the daily data (*χ*^2^/d.o.f.). Which model provides us with better understanding of epidemics? Overall, the values of the scaled RMS’s and the scaled (*χ*^2^/d.o.f)’s in Tables 1 & 2 suggest that the ALM is superior. However, examination of the figures shows that these differences are largely due to the declining phases of the daily curves, and that the DLM provides a good fit to the initial rises and the linear phases of the curves. Thus the DLM will be used to estimate the parameter *T* and the ALM to determine the parameter *S* (see §7).

The next two sections show the cases and deaths data for Italy and the U.K., and display the best fits to the DLM and the ALM for each data set.

### 6.3. The Epidemic in Italy

Figures 6 & 7 show the fits of the DLM and the ALM to the histories of cases and deaths in Italy. The parameters of the fits are displayed in Tables 1 & 2.

**Fig. 6.**
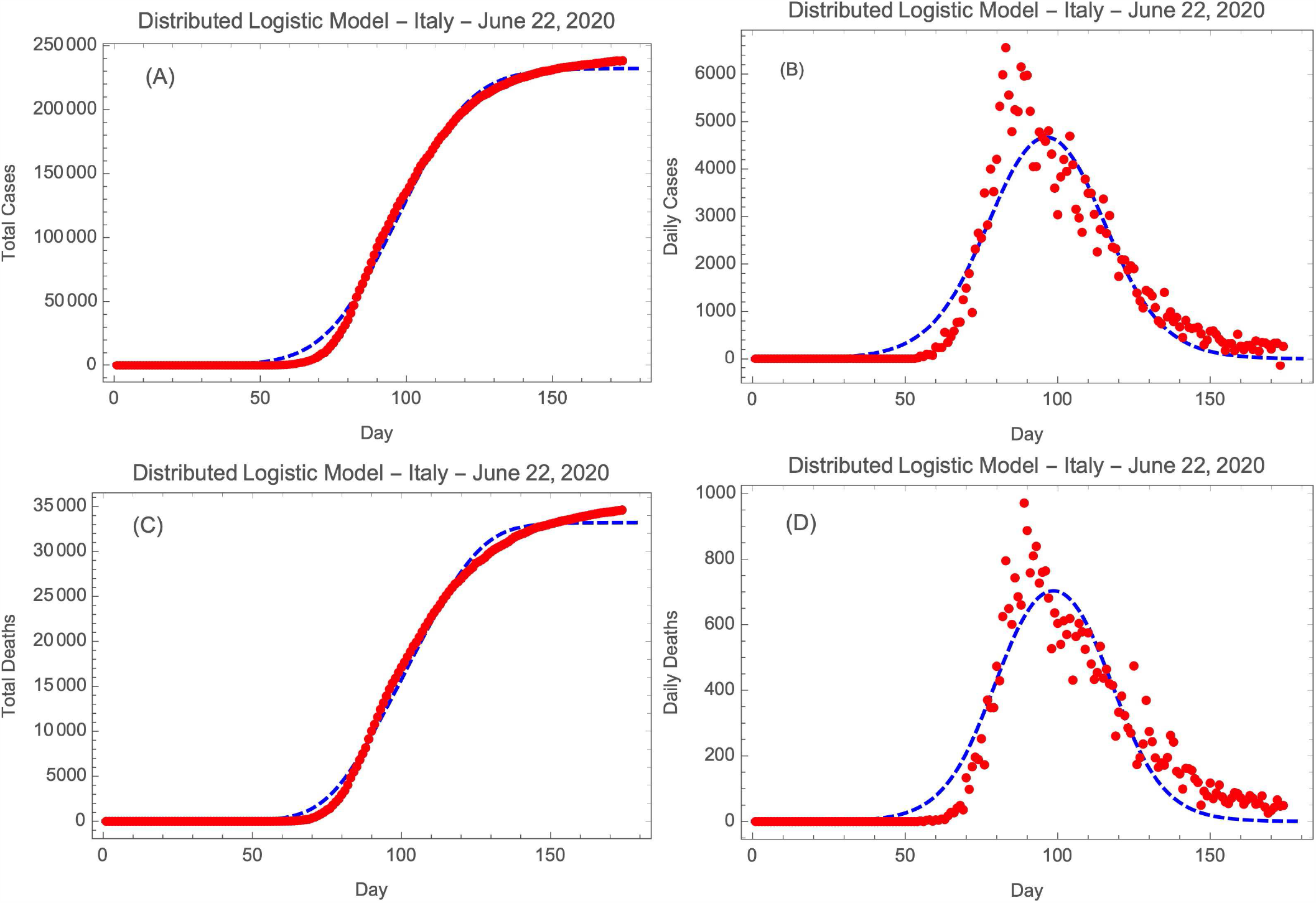
DLM fits to (A) total cases, (B) daily cases, (C) total deaths, and (D) daily deaths in Italy. The points are the data and the curves are the fits to Eqns. 5 & 6.

**Fig. 7.**
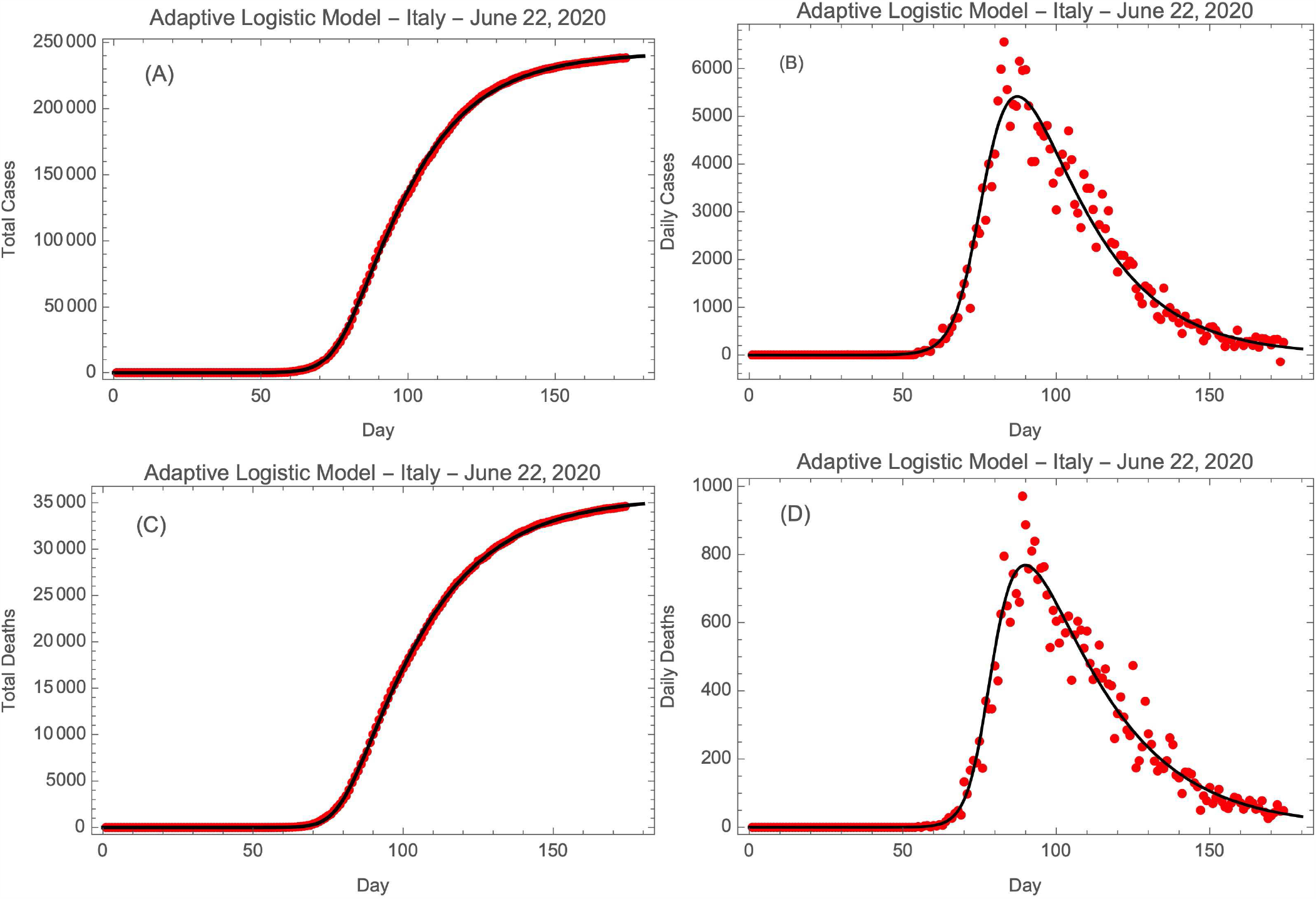
ALM fits to (A) total cases, (B) daily cases, (C) total deaths, and (D) daily deaths in Italy. The points are the data, the total numbers curves are the fits to Eqn. 7, whereas the daily numbers curves are their derivatives.

### 6.4. The Epidemic in the United Kingdom

Figures 8 & 9 show the fits of the DLM and the ALM to the histories of cases and deaths in the U.K. The parameters of the fits are displayed in Tables 1 & 2.

**Fig. 8.**
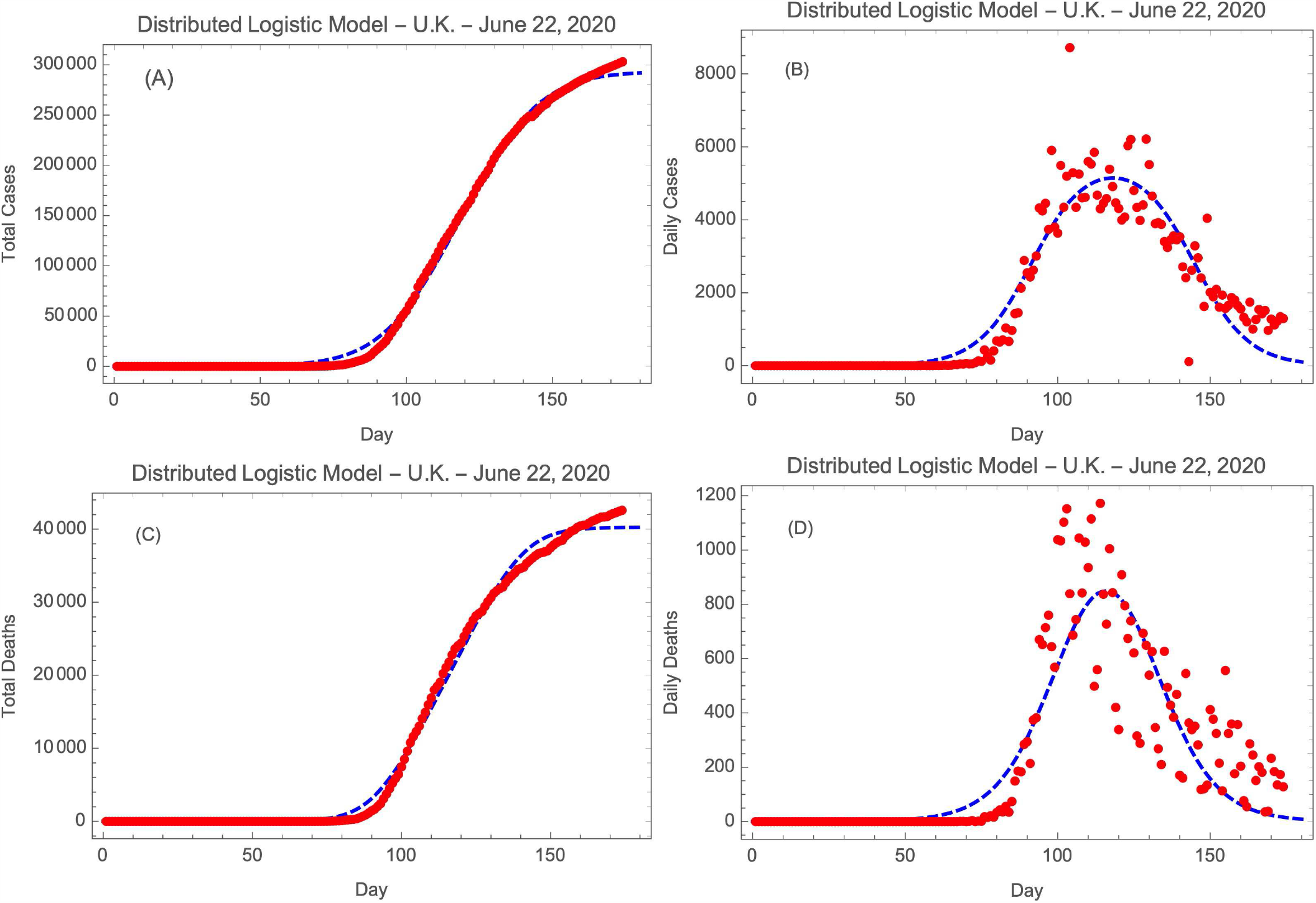
DLM fits to (A) total cases, (B) daily cases, (C) total deaths, and (D) daily deaths in the U.K. The points are the data and the curves are the fits to Eqns. 5 & 6.

**Fig. 9.**
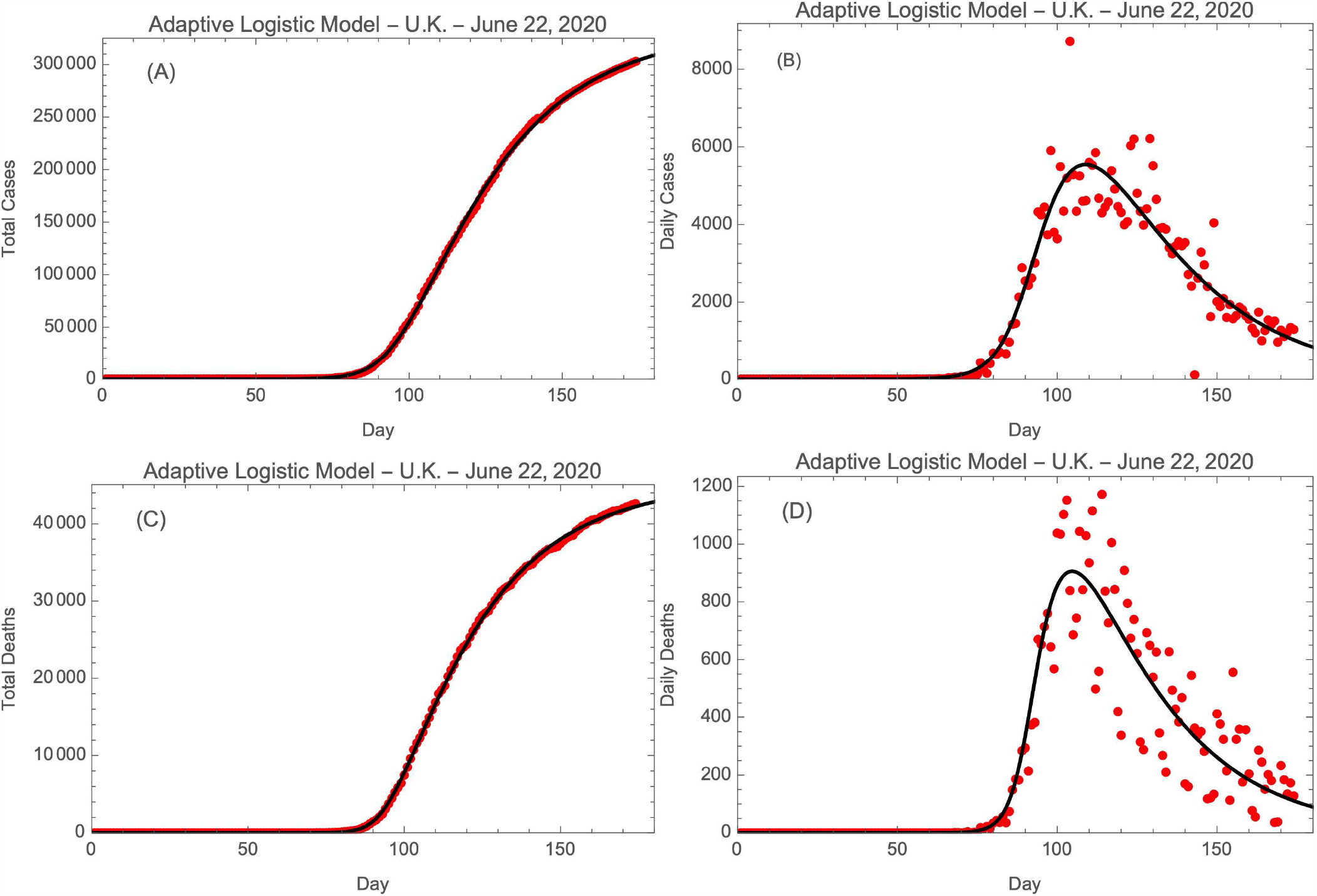
ALM fits to (A) total cases, (B) daily cases, (C) total deaths, and (D) daily deaths in the U.K. The points are the data, the total numbers curves are the fits to Eqn. 7, whereas the daily numbers curves are their derivatives.

## 7. Discussion

### 7.1. The History of Cases Versus that of Deaths

The history of testing in the U.S. is such that early on in the epidemic cases were recognized only when people exhibiting symptoms were tested, but as testing became more widespread, cases were discovered in asymptomatic individuals. This has the effect of increasing the number of cases as the epidemic proceeds. The same is not true for deaths, so one expects deaths to provide a more accurate measure of the history of the epidemic. Indeed, examination of Figures 4, 7 & 9 shows a faster decline in the number of deaths than in the number of cases, as expected from this argument. The parameters derived from the deaths data will be adopted in the analysis below.

### 7.2. The Mortality Ratios and Infection Delays

It is interesting to learn if and how the mortality ratio *M* evolved during the epidemics in each country. One analysis found the best single *M* and time shift Δ*t* for each country by shifting and scaling the raw daily cases and comparing the results to the deaths data; it found *M* and Δ*t* by minimizing the summed squares of their differences. Fig. 10 shows the shifted and scaled data, and the best fit parameters *M* and Δ*t* are in Table 3. The ratio *M* was found to be significantly lower in the U.S. than in Italy or the U.K. The delays between the cases and deaths curves were about 6 days for the U.S. and Italy, but zero for the U.K. (which we do not understand). The delays presumably represents the average course of the disease for each fatality, confounded by differences in reporting of the two phenomena.

**Table 3.** Mortality Ratios and Time Delays.

| Parameter | U.S. | Italy | U.K. |
| --- | --- | --- | --- |
| $N_{\text{deaths}}/N_{\text{cases}}$ | 0.0531 | 0.145 | 0.141 |
| Mortality ratio $M$ | 0.061 | 0.15 | 0.15 |
| Delay $\Delta t$ (days) | 7 | 6 | 0 |

**Table 4.** Adopted Parameters (from Deaths Data)

| Parameter | Model. | U.S. | Italy | U.K. | Mean |
| --- | --- | --- | --- | --- | --- |
| $N_{\text{data}}$ | ... | 174 | 174 | 174 | ... |
| $N_{\text{cases}}$ | ... | 2,255,119 | 238,275 | 303,310 | ... |
| $N_{\text{deaths}}$ | ... | 119,719 | 34,610 | 42,589 | ... |
| $r$ (days<sup>-1</sup>) | BLM<sup>a</sup> | 0.070 | 0.085 | 0.088 | $0.081 \pm 0.008$ |
| $r$ (days<sup>-1</sup>) | DLM<sup>a</sup> | 0.106 | 0.116 | 0.100 | $0.107 \pm 0.08$ |
| $r_0$ (days<sup>-1</sup>) | ALM<sup>b</sup> | 0.287 | 0.284 | 0.288 | $0.286 \pm 0.002$ |
| $T$ (days) | DLM<sup>a</sup> | 51. | 37. | 29. | $39. \pm 11$ |
| $S$ | ALM<sup>b</sup> | 15,300 | 6,040 | 6,650 | ... |

<sup>a</sup>Estimated from daily data.
<sup>b</sup>Estimated from total data.

**Fig. 10.**
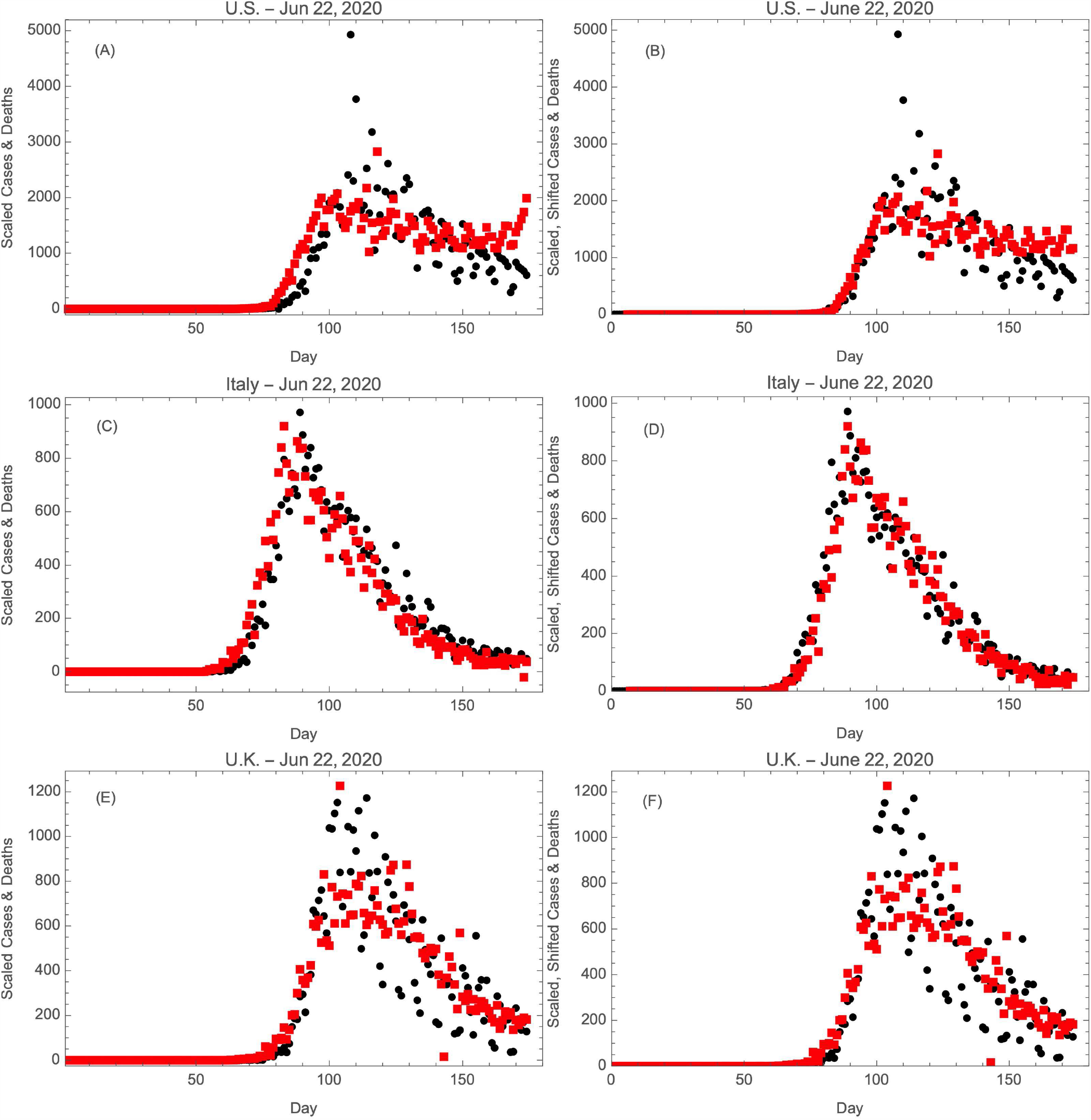
Deaths (black circles) and scaled cases (red squares) in (A) the U.S., (C) Italy, and (E) the U.K. Deaths (black circles) and scaled and shifted cases (red squares) in (B) the U.S., (D) Italy, and (F) the U.K. These should be compared with Figs. 4, 7, & 9.

Second, Fig. 11 shows the instantaneous mortality ratios calculated by dividing the deaths by cases (shifted by Δ*t*); it also shows the mean mortality ratios *M* just determined. A general trend is that the mortality ratios vary slowly with time in each country.

**Fig. 11.**
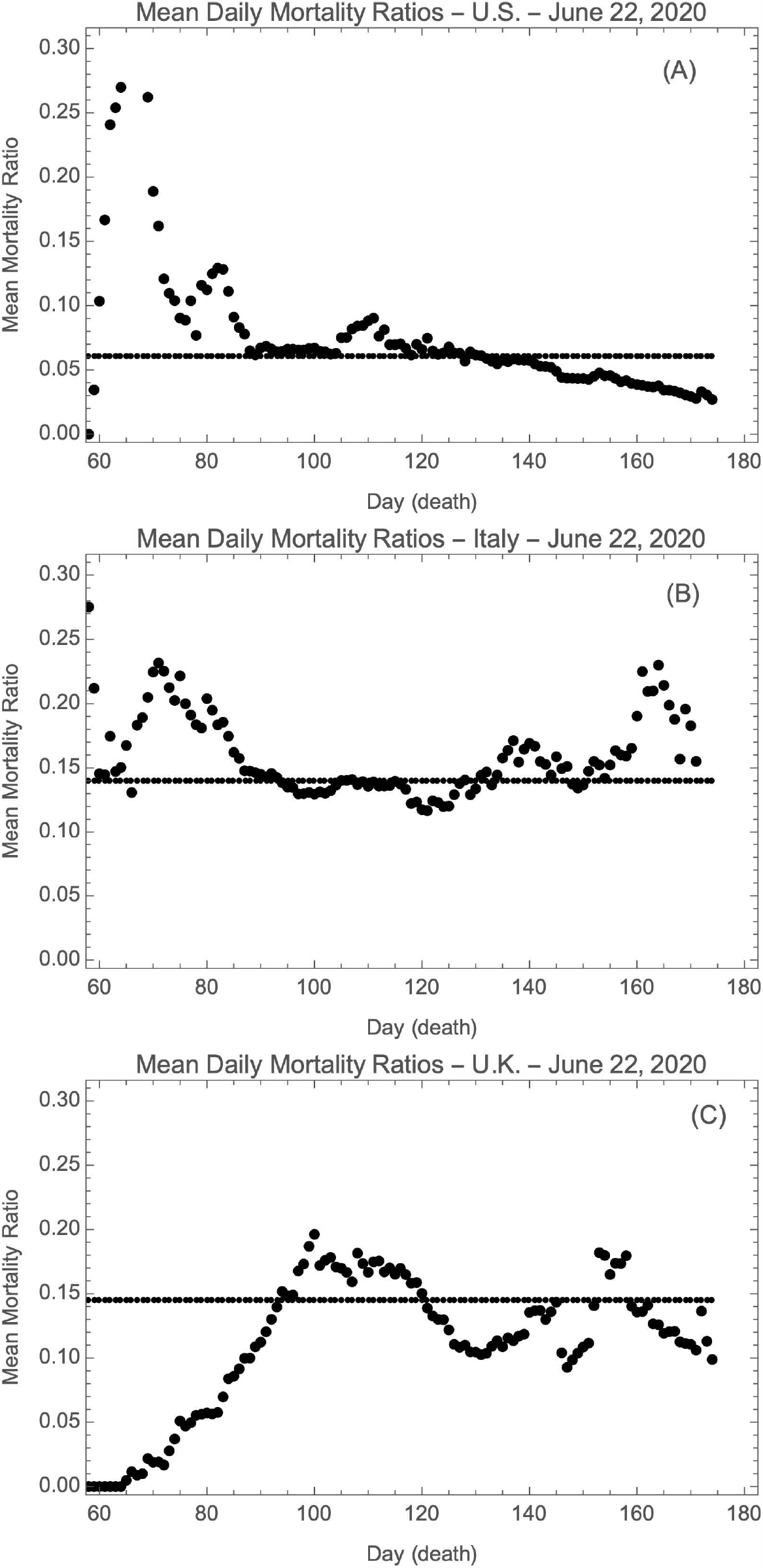
Day by day mortality ratios (points) and mean mortality ratio (black line) in (A) the U.S., (B) Italy, and (C) the U.K. (using seven day running means).

**Fig. 12.**
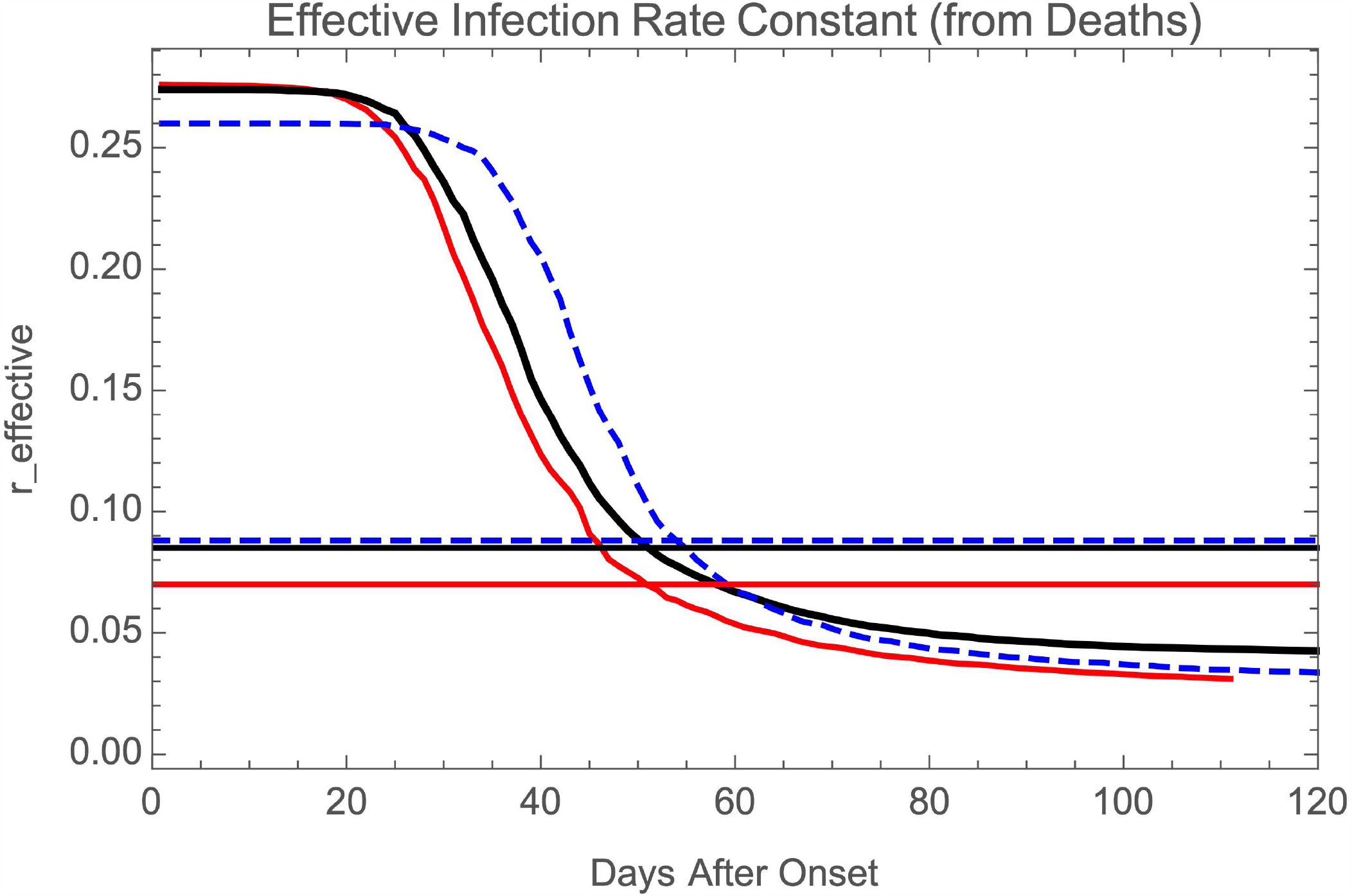
Time behavior for the effective infection rate constant in the ALM as a function of the number of days since the onset of each epidemic, for the U.S (red curves), Italy (black curves), and the U.K. (dashed blue curves). The horizontal lines are *r*’s derived using the BLM and the daily deaths data.

### 7.3. Comparison of Parameters in Common in DLM and ALM

1. For Italy and the U.K., in both the DLM and the ALM the *K*’s closely equal the total numbers *N*_tot_ (sizes of the data sets). However for the U.S. *K* is significantly larger than *N*_tot_, suggesting that the epidemic has more time to run.
2. For each model the initial infection rate constants are comparable in the three countries, and they are different for the DLM (0.11) and ALM (0.29). This difference is because *r* and *r*_0_ have different meanings in the DLM and the ALM.
3. It is important to realize that the parameter *r*, the infection rate constant in the BLM and DLM, is distinct from the initial infection rate constant *r*_0_ of the ALM. While *r* is truly a constant, *r*_0_ is the infection rate constant only at the outset of the epidemic; it is the effective infection rate constant *r*_eff_ that determines the rate *dg/dt* at any later time. Eqn. 8 shows how this goes, and it’s illustrated in Fig. 12, which compares the effective values *r*_eff_ (*t*) along with the values of *r* from the BLM (Table 4) for each country. Since the way *r*_eff_ varies with time depends on the value of *S*, which is different for each country, a simple comparison of *r*_0_’s across countries would be misleading. The correct treatment is to compare plots of *r*_eff_ (*t*) as each epidemic unfolds, as shown in Fig. 12. From these plots one can conclude that the effective infection rate constants for all three countries are essentially identical for given times after the onset of each epidemic.
4. As discussed above, one expects the behavior of the numbers of deaths with time to more accurately reflect the course of the epidemics than does the behavior of cases. Because the fits to the ALM are systematically superior to those to the DLM, the ALM values were adopted in each case when they were available. With these things in mind, Table 4 presents the best estimates of the properties of the epidemics, listed by country.

## 8. Results

A summary of the results of this paper is:

1. Models with fixed infection rate constants *r* (the BLM and DLM) do not provide good representations of the full course of the epidemic in any country.
2. Comparing Figs. 1 and 2, one can see the effect of adding the time smoothing parameter *T* on the DLM curves. The result fits much better the widths of the data history, and better fits the rise of the epidemics. The result is *T* (US) *> T* (Italy) *> T* (UK). This is what is expected given the geographical sizes of the three countries. The value *T* (US) ≃ 50 days is comparable to the interval over which hot spots developed across the U.S. during the spring of 2020 (New York Times, 2020). One expects that subsequent analysis with the DLM will find *T* to have increased as more and more hot spots arise due to the easing of mitigating factors across the countries, especially in the U.S.
3. The ALM can well describe the epidemics, and its best estimates of the effective infection rate constants *r*_eff_ as a function of time show them to be essentially identical for all three countries.
4. For each model separately the initial infection rate constants are essentially identical in the three countries.
5. The fact that the effective infection rate constant declines with time is likely the product of increasing mitigation efforts and improving medical treatments.
6. The time-averaged mortality ratio in the U.S. was about 0.061, while in Italy and U.K. it was much higher at about 0.14. In all three countries it tended to vary slowly through each epidemic.

## 9. Concluding Remarks

This paper has presented two new versions of the logistic model that describe the evolution of the number of infections in an epidemic as a function of time. The first modification, the Distributed Logistic Model (DLM), approximates the spread of disease centers across a large country like the United States; this introduces a single new parameter *T* that measures the time span over which different hotspots appear. It is successful in capturing the initial and linear phases of the epidemics in each country. However, it is intrinsically time-symmetric, with the rising and declining phases of the daily curves being mirror images, which does not match the data. The basis of the second modification, the Adaptive Logistic Model (ALM), is an infection rate constant that declines with time as public health and social mitigation measures are introduced; this requires a single new parameter *S* that parameterizes the point at which social mitigation efforts take effect. This model, which is intrinsically asymmetric across an epidemic, is especially successful at capturing the declining phases of epidemics. When fit to all of the data for number of cases and number of deaths, the results for both models are good, especially for the ALM. The results of the fitting show (1) that it took about two months for the epidemic to spread across the U.S., and somewhat less time for Italy and the U.K, and (2) that mitigation efforts have been equally successful in the three countries.

These new approaches are useful ways to understand the histories of epidemics. In particular, each provides a quantitative measure of a parameter that helps to control the evolution of an epidemic. However, they neither are intended to replace traditional epidemiological analysis, nor are they suitable for predicting the future, for example, in the event that mitigation efforts are significantly modified.

## Data Availability

All data and code used in this study are available to any person.

## 10. Acknowledgements

Brian Boyle, Mary Roberts, and Bob Sauer provided very helpful comments and suggestions.

## Footnotes

2 This is a version of the Bernoulli differential equation *f* ^′^ = *f* (1 − *f*).

3 Fitting the daily numbers is statistically the preferred procedure as the data points are independent, unlike those for daily totals. Where there are results from fitting total data and from fitting daily data, the latter will be adopted.

4 The use of the logistic equation to fit the data on deaths is justified in below.

5 The DLM was introduced by Roberts (2020) who analyzed only U.S. cases.

6 Models where the function is of the form *e*^−g/S^ or *e*^−t/τ^, where *τ* is a characteristic time, were also explored, as were hybrid models that included both factors. None of these provided fits to the data as close as the choice 1*/*(1 + *g/S*), so it was used in Eqn. 7.

7 Eqn. 7 is so named because it models a population’s adaptation to the disease.

8 A complete Mathematica™ notebook that performs the solution is available from the author.

9 Eqn. 7 cannot be used to fit directly the daily numbers.

10 Section 7.2 below shows that the mortality ratio evolves only slightly through each epidemic.

11 The values of the infection rate constant *r* derived from the BLM will be used to compare to the results from the DLM and the ALM.

12 To account for the larger numbers of cases than deaths, the values for the cases data were scaled by the mean mortality ratio to compare them with the results for the death data.

13 Poisson statistics were used for the uncertainties of each datum, where 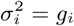, where *g* is the model prediction for the i^th^ datum. This leads to *χ*^2^ per degree of freedom greatly in excess of unity, suggesting that this estimate for *σ*i is significantly too small. Although these statistics cannot be used to determine a goodness of fit, they can be used to compare qualitatively the fits for different models and data sets.

## References

AHA (2020), American Hospital Association, “A Compendium of models that predict the spread of COVID-19,” https://www.aha.org/guidesreports/2020-04-09-compendium-models-predict-spread-covid-19.

Batista, M. (2020), “Estimation of the final size of the coronavirus epidemic by the logistic model (Update 4),” https://www.researchgate.net/publication/339240777.

Best, R., & Boice, J. (2020), “Where The Latest COVID-19 Models Think We’re Headed– And Why They Disagree,” https://projects.fivethirtyeight.com/covid-forecasts/.

CDC (2020), Centers for Disease Control and Prevention, https://www.cdc.gov/coronavirus/2019-ncov/covid-data/forecasting-us.html.

Chen, Y.-C.,Lu, P.E., Change, C.H., & Liu, T.H. (2020), “A Time-dependent SIR model for COVID-19 with Undetectable Infected Persons,” http://gibbs1.ee.nthu.edu.tw/ A TIME DEPENDENT SIR MODEL FOR COVID 19.PDF.

Chen, D-G., Chen, X., & Chen, J.K. (2020), “Reconstructing and forecasting the COVID- 19 epidemic in the United States using a 5-parameter logistic growth model,” Global Health Research and Policy 5:25.

ECDPC (2020), European Centre for Disease Prevention and Control, https://www.ecdc.europa.eu/en/publicdata/download-todays-data-geographic-distribution-covid-19-cases-worldwide (2020), data retrieved June 22, 2020.

Hethcote, H.M. (2000) “The Mathematics of Infectious Diseases,” SIAM Review, 42, 599–653.

Huang Y, Yang L, Dai H, Tian F & Chen K. Epidemic situation and forecasting of COVID- 19 in and outside China (2020), [Preprint]. Bull World Health Organ. E-pub: 16 March 2020. doi:http://dx.doi.org/10.2471/BLT.20.255158

Kermack, W.O., & McKendrick, A.G. (1927), “A Contribution to the Mathematical Theory of Epidemics,” Proc R Soc Lond A, 115, pp. 700–721.

Lavrova, A.I., Postnikov, E.B., Manicheva, O.A., & Vishnevsky, B.I. (2017), “Bi-logistic model for disease dynamics caused by mycobacterium tuberculosis in Russia,” R Soc Open Sci, 4, 171033.

New York Times (2020), multiple news stories (New York, New York).

Roberts, D. H. (2020), “The Distributed Logistic Model for Pandemics. I. Application to the COVID-19 Pandemic of 2020 in the United States,” http://preprints.org/10.20944/preprints202005.0395.v1.

Shen, C. Y. (2020), “Logistic growth modelling of COVID-19 proliferation in China and its international implications,” International Journal of Infectious Diseases, 96, 582–589.

Verhulst, P.-F. (1838). “Notice sur la loi que la population poursuit dans son accroissement,” Correspondance Mathématique et Physique, 10, 113–121.

Wangping, J., Ke, H., & Yang, S. et al. (2020), “Extended SIR Prediction of the Epidemics Trend of COVID-19 in Italy and Compared With Hunan, China, “ Front Med (Lausanne). 2020;7:169. Published 2020 May 6. doi:10.3389/fmed.2020.00169.

Wu, K., Darcet, D., Wang, Q. & Sornette, D. (2020), “Generalized logistic growth modeling of the COVID-19 outbreak in 29 provinces in China and in the rest of the world,” preprint (2003.05681).

Zhao, S., & Chen, H. (2020), “Modeling the Epidemic Dynamics and Control of COVID- 19 Outbreak in China,” https://www.researchgate.net/publication/339597236.

